# Adiposity and Sex Influence on SARS-CoV-2 Antibody Response in University Students. An ESFUERSO cross-sectional study

**DOI:** 10.1101/2023.12.15.23298521

**Authors:** Adriana L. Perales-Torres, Lucia M. Perez-Navaro, Esperanza M. Garcia-Oropesa, Alvaro Diaz-Badillo, Yoscelina Estrella Martinez-Lopez, Marisol Rosas, Octelina Castillo, Laura Ramirez-Quintanilla, Jacquelynne Cervantes, Edda Sciutto, Claudia X. Munguia Cisneros, Carlos Ramirez-Pfeifer, Leonel Vela, Beatriz Tapia, Juan C. Lopez-Alvarenga

## Abstract

**Introduction:** Prior studies have identified various determinants of differential immune responses to COVID-19. This investigation delves into the Ig-G anti-RBD marker, scrutinizing its potential correlations with sex, vaccine type, body fat percentage, metabolic risk, perceived stress, and previous COVID-19 exposure.

**Methods:** In this study, data were obtained from 116 participants from the ESFUERSO cohort, who completed questionnaires detailing their COVID-19 experiences and stress levels assessed through the SISCO scale. Quantification of Ig-G anti-RBD concentrations was executed using an ELISA assay developed by UNAM. Multiple regression analysis was adeptly employed to control for covariates, including sex, age, body fat percentage, BMI, and perceived stress.

**Results:** This sample comprised young individuals (average age of 21.4 years), primarily consisting of females (70%), with a substantial proportion reporting a family history of diabetes, hypertension, or obesity. Most students had received the Moderna or Pfizer vaccines, and 91% displayed a positive anti-RBD response.

A noteworthy finding was the interaction between body fat percentage and sex. In males, increased adiposity was associated with a decrease in Ig-G anti-RBD concentration, while in females, the response increased. Importantly, this trend was consistent regardless of the vaccine received. No significant associations were observed for variables such as dietary habits or perceived stress.

**Conclusions:** In summation, this research reports the impact of both sex and body fat percentage on the immune response through Ig-G anti-RBD levels to COVID-19 vaccines. The implications of these findings offers a foundation for educational initiatives and the formulation of preventive policies aimed at mitigating health disparities.

## Introduction

To understand factors that can impact the serum levels of antibodies produced by the COVID-19 vaccine, many studies have been conducted. Age is a critical factor that plays a significant role in determining the immune response. Elderly individuals with obesity and non-prior infection had reduced antibody titers against SARS-CoV-2 spike antigen after CoronaVac vaccine (manufactured in China) compared to non-obese people.(1) Lower antibody response, after receiving two doses of the Pfizer vaccine, has also been linked to central obesity (correlation of r= -0.3), presence of hypertension and smoking habits, with no notable differences by gender.(2)

Interestingly, losing weight has been shown to improve the adaptive immune response particularly an increase in INF-g2 levels following administration of two doses of mRNA vaccine.(3)

Sex and body weight interaction can also result in varying immune responses. For individuals with a BMI >40 kg/m2, there were no discernible differences in IgG antibody levels between sexes. In contrast, those with normal weight showed higher levels among males.(4)

The present study focused on a nested sample of students from the ESFUERSO program, which recruited first-year college students living in Reynosa prior the COVID-19 pandemic in 2018. This group of students reported a significant family history of type 2 diabetes (T2D) with 25% having family history of the condition, and 39% having family history of hypertension. Interestingly, between 17 to 47% students were unsure whether their first-degree relatives suffered from T2D, hypertension, or other obesity-related metabolic issues, as documented in the ESFUERSO study.

These young students experienced impact from the COVID-19 pandemic, forcing them to home confinement, which led to changes in their food intake, physical activity, and increased psychological stress. Vaccination efforts began in 2021, employing novel mRNA vaccine used in veterinary science for three decades.

During the pandemic, it was evident that metabolic imbalances associated with obesity could increase the severity of COVID-19 and the risk of mortality.(5, 6) The aim of this study was to analyze the immune response to the receptor-binding domain (RBD), a protective epitope found in the S protein, using Ig-G response among young students from Mexico, living near the US-Mexico border. The study provides insights in the immune response and the potential implications in the context of COVID-19.

## Methods

### Study Sample

In 2018, the ESFUERSO (Estudio en la Frontera Urbana para las Enfermedades y Factores Asociados a la Obesidad) cohort study was launched, with its focus on first-year students from two universities in Reynosa, Tamaulipas. Initially, the parental sample included 500 students. However, due to the pandemic, a subset of 116 students was contacted between September 1^st^ and October 31^st^, 2021. During this period, we obtained signed informed consent (see Ethics Statement below for details), conducted questionnaire surveys, collected anthropometric measurements, and collected blood samples from 108 students.

### Measures

The questionnaires and methods used in the ESFUERSO study have been described elsewhere.(7) Briefly, the questionnaires collected information on family metabolic risk, anxiety, depression and uncertainty. The Cronbach α coefficient from each question ranged from 0.72–0.96. The test-retest for agreement in categorical variables was a kappa coefficient between 0.5–0.91 and an intraclass correlation coefficient between 0.73–0.96 in continuous variables. The stress during the pandemic was evaluated with the Coping Inventory for Stressful Situations (SISCO), composed of five stressors, five symptoms and five coping strategies. The SISCO had a Cronbach α coefficient of 0.9 with high homogeneity and was validated in Mexico and other Latin American countries.(8) All the questionnaires were administered electronically, with students completing them on their own cellphones. This approach ensured efficient data collection and minimized the need for physical paperwork or in-person administration. Weight, height, acanthosis nigricans grade(9) were assessed and registered at the universities by standardized nutritionist.(7) Blood samples were obtained after an overnight fasting period for the measurement of serum concentration of Ig-G anti-RBD by indirect ELISA.(10) Following collections, the samples underwent centrifugation, and four aliquots were stored at -20 C. These aliquots were subsequent transported to Mexico City in November 2021 for antibody analysis. The assessment of the effective neutralizing concentration of anti-RBD IgG was carried out, and this variable was analyzed in both continuous and dichotomous dimensions. A threshold of 1.0 for anti-RBD IgG ratio was established to differenciate between effective and non-effective neutralization.

### Ethics Statement

The protocol and informed consent were approved by the Comite de Etica Institucional de la Unidad Academica Multidisciplinaria Reynosa-Aztlan (CEI-UAMRA) number registration CEI-UAMRA 005/2019/CEI under Health normativity (NOM-012-SSA-3-212). All participants signed the approved informed consent. The present report followed the STROBE recommendations for cross-sectional studies.(11)

### Statistical analysis

Descriptive statistics with percentage for counts variables, mean and standard deviation for continuous variables. Inferential statistics with regressions was used for the SISCO questionnaire for anxiety, uncertainty, sadness, lack of sleep was contrasted by the presence of metabolic risk. Locally weighted regression (lowess) was performed to analyze nonlinear functions. We used the antibody concentration as dependent variable adjusted for sex, age, metabolic risk, body fat percentage and BMI. Multiplicative interactions for covariates were analyzed. Variance inflation factors (VIF) was calculated to evaluate multicollinearity for lineal models without interactions. The models were evaluated using first to third grades polynomial multiple regression and goodness of fit with mean squared error calculation. The best goodness of fit for data was polynomial multiple regression with 114 students with all complete data. All analyses were performed with Stata V18.0 (StataCorp, College Station, TX).

## Results

A total of 108 students in the ESFUERSO cohort were enrolled during the 3rd year of the cohort follow-up in Reynosa. The participants had mean age of 21.4 (SD 1.0) years, average BMI of 27.9 (SD 6.2), and gender distribution of 69% (75 out of 108) females. A significant family risk of T2D, hypertension, presence of obesity, or a combination of these conditions was identified in 70% of the students. Notably, there were no discernible differences by universities on metabolic risk, anthropometry, sex, and commercial brand of vaccine or presence of adequate antibody levels. Only 3 (3%) students from the private university had no COVID-19 vaccination at the time of the study and they also belonged to the metabolic risk group (Pearson standardized distance >3.0). From vaccinated students 97 (90%) were vaccinated with Moderna or Pfizer and only 8 (7%) with other vaccines (Johnson & Johnson n=1, Cansino n=6, Sinovac n=1). The prevalence of positive anti-RBD was 91%.

**Table 1.** Descriptive Statistics of General Variables by Sex. Continuous variables are presented as mean and standard deviation. Ordinal variables, such as the perceived stress items of the SISCO questionnaire, are described using a 6-point Likert scale (0 = none, 5 = highest score). Acanthosis nigricans is described by its median and interquartile range (Q1, Q3).

| Variable | Females (n= 75) | Males (n= 33) |
| --- | --- | --- |
| Continuous variables [Average (SD)] |  |  |
| Age (years) | 21.4 (1.1) | 21.4 (0.9) |
| BMI (kg/m<sup>2</sup>) | 27.7 (6.1) | 28.4 (6.3) |
| Body fat (%) | 33.04 (10.45) | 24.2 (10.34) |
| Waist circumference (cm) | 83.3 (13.2) | 91.9 (13.7) |
| Neck circumference (cm) | 34.4 (3.2) | 40.4 (10.6) |
| Systolic blood pressure (mmHg) | 113 (9) | 120.8 (18.7) |
| Diastolic blood pressure (mmHg) | 77 (7) | 81.1 (92.9) |
| Ordinal variables [median (Q1, Q3)] |  |  |
| Anguish | 3 (2, 5) | 3 (1, 4) |
| Uncertainty | 3 (2, 4) | 2 (1, 4) |
| Lack of sleep | 4 (2, 5) | 2 (1, 3) |
| Sadness | 3 (3, 5) | 2 (1, 3.5) |
| Anxiety | 4 (3, 5) | 3 (1.5, 4) |
| Acanthosis nigricans | 0 (0, 2) | 1 (0, 2) |

The multicollinearity analysis showed BMI and fat percentage had VIF= 4.8 and 1/VIF= 0.02, most models were analyzed separating both variables. The body fat percentage interaction with sex was statistically significant, explaining why the serum concentration of anti-RBD decreased as adiposity increased in men (p=0.034 for second grade term), but Ab-RBD increased with increased adiposity in women (p=0.01, p=0.015 for second and third grade terms) (Figure 1). The interaction remained in spite of the type of vaccine. The adjusted model minimized the mean squared error (0.51 and R^2= 0.14), compared with other models (MSE between 0.54 to 0.56, and R^2 between 0.04 and 0.07) (Figure 2). The residuals adjusted a normal distribution (Shapiro-Wilkins p=0.136).

**Figure 1.**
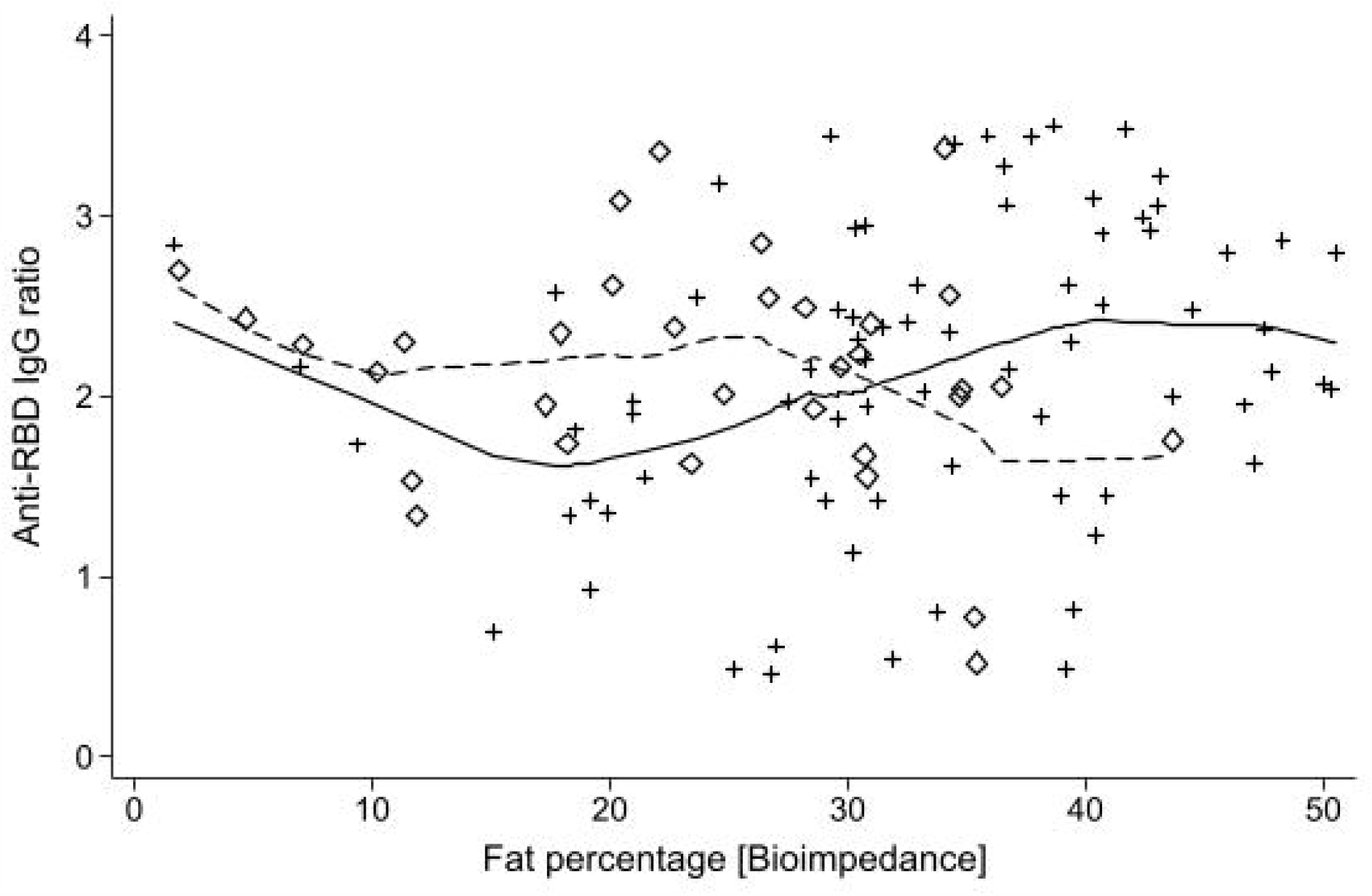
Sex differences analyzed with locally weighted regression (lowess) function of neutralizing anti-RBD IgG ratio and body fat percentage in a sample of university students. Females: Diamonds, Males: cross, Lowess females: Continuous line, Lowess males: discontinuous line.

**Figure 2.**
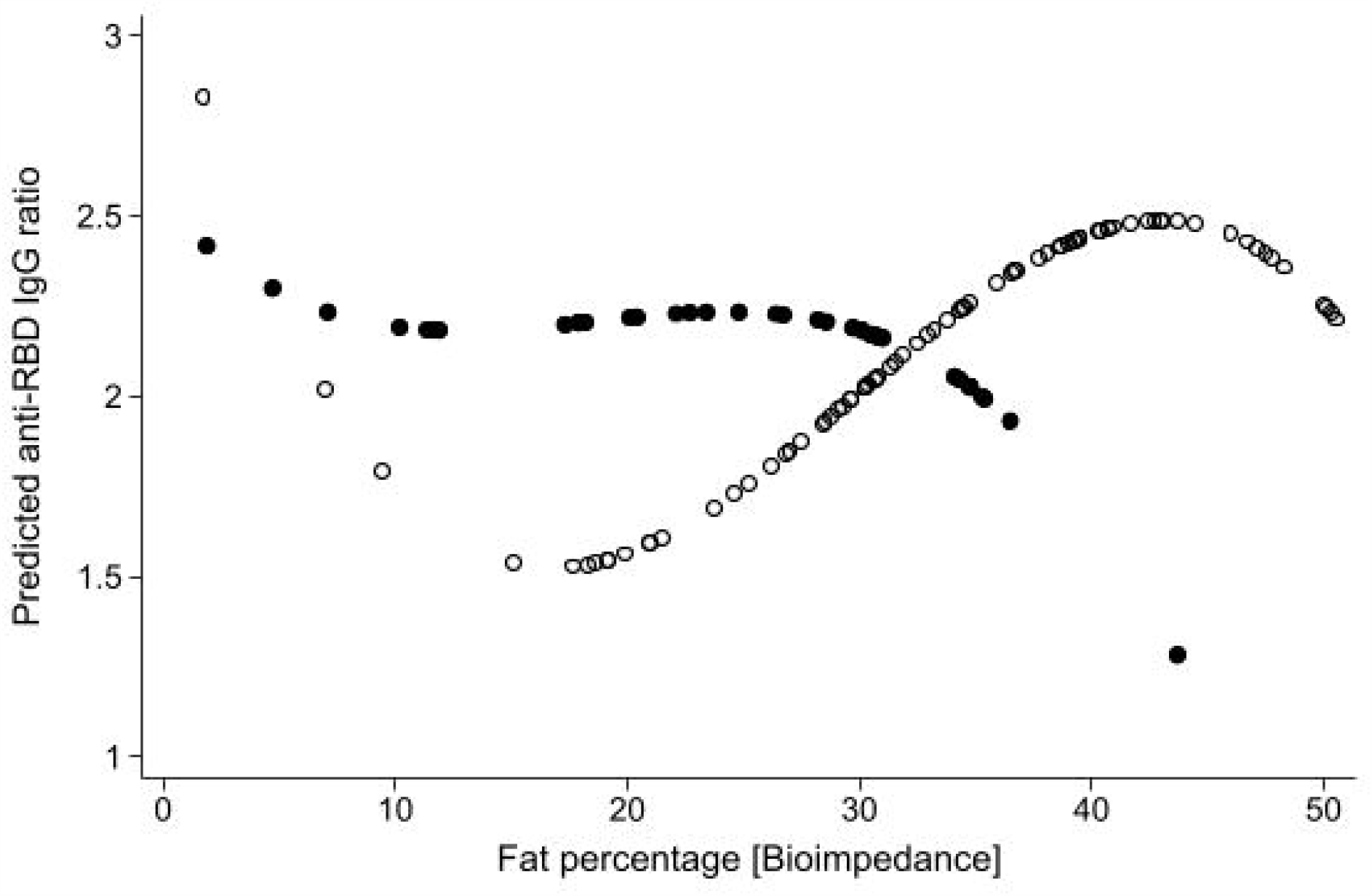
Function of anti-RBD IdG ratio predicted by body fat percentage with a 3^rd^ grade polynomial regression model. The function for males can be modeled with a 2^nd^ grade polynomial. Females: White circles, Males: Black circles.

No differences due to metabolic risk factors or effective antibody concentration were found, for food consumption, distress, and uncertainty, lack of sleep, sadness, and anxiety.

**Figure 1.** Sex Differences Analyzed with Locally Weighted Regression (Lowess) Function. This figure displays the relationship between neutralizing anti-RBD IgG ratio and body fat percentage in a sample of university students. Females are represented by diamonds, and males by crosses. The Lowess curve for females is shown as a continuous line, while for males, it is depicted as a discontinuous line.

**Figure 2.** Adjusted Function of Anti-RBD IgG Ratio Predicted by Body Fat Percentage. This figure illustrates the relationship between anti-RBD IgG ratio and body fat percentage using a 3rd-grade polynomial regression model. For males, a 2nd-grade polynomial is sufficient. Females are represented by white circles, and males by black circles.

## Discussion

Based on the results of our study, it appears that the neutralizing anti-RBD response to the COVID-19 vaccine is influenced by a multiplicative interaction of sex and body fat percentage. Specifically, females tend to have increased responses while males tend to have decreased responses (Figure 2). Stress scores do not appear to have significant effects.

This observation aligns with existing research on sex-based differences in immune responses. For instance, a study involving the Cameron County Hispanic Cohort, which included 624 participants with a mean age of 50 (SD 14) years, has previously reported sex-specific variations in adipokines and carotid intima media thickness.(12) The present study extends these findings to a younger cohort, specifically individuals in the final stages of adolescence residing near the U.S.-Mexico border. This highlights the relevance of considering age and geographic location when examining immune response differences between sexes.

Additionally, another study conducted in Mexico on 980 adult participants with a median age of 50 (Q1: 36, Q3: 54) who had obesity before mass vaccination sheds light on this matter. (13) The authors identified independent factors associated with SARS-Cov-2 infection in a symptomatic group. Their findings revealed higher levels of anti-S1/2 antibodies in individuals with conditions like advanced age, type 2 diabetes, hypertension, and a positive correlation with body mass index (BMI). Furthermore, women exhibited higher levels of anti-RBD IgG antibodies compared to men. This study highlighted the heightened vulnerability of individuals with underlying health conditions or obesity to SARS-CoV-2 infection. In contrast, the ESFUERSO cohort focused on a younger population who received vaccinations, providing valuable insights into this demographic group.

Other populations have reported similar findings. Yamamoto et al.(14), reported sex– associated differences in the relationship between body mass index and SARS-CoV-2 antibody titers following the BNT162b2 vaccine in a study of 2,435 healthcare workers in Japan. Additionally, a meta-analysis examining antibody responses to COVID-19 vaccinations also indicated a significant association between obesity and reduced antibody response. (15) Nevertheless, the considerable heterogeneity (88%) observed across studies suggests that biological factors, including sex, age, and body fat play a pivotal role in this outcomes.

Tailoring vaccination plans based on an individual’s characteristics may enhance vaccine effectiveness. Addressing gender-specific and body fat-related factors in public health interventions have the potential to reduce infection rates. Using the provided information in this study can help in programs to educate individuals about their susceptibility to infections considering the social determinants in the U.S.-Mexico border region.

## Limitations

While the present study yields valuable insights, it is important to acknowledge several potential limitations. Variations in immune responses across different age groups, the influence of genetic factors, and the impact of social determinants can introduce complexities that our study may not fully capture. Moreover, it’s essential to recognize that the study’s cross-sectional design allows identifying associations but does not establish causality. These considerations emphasize the need for caution in generalizing the findings and highlight avenues for further research.

In summary, this study provides novel insights into the response of anti-RBD IgG antibodies to vaccination in a young cohort. The findings reveal a complex relationship between sex and body fat percentage, depicted by a third-degree polynomial curve (Figure 2). This emphasizes the intricate interplay between body fat and the immune response to vaccines and accentuates the importance of considering sex-specific factors, especially among younger individuals. Comprehensive knowledge of distinct characteristics and immunological responses, help to understand social and biological dynamics for tailoring vaccination strategies, optimizing public health interventions and reducing health disparities

## Data Availability

Data will be available in the UTRGV repository open as CC BY. https://scholarworks.utrgv.edu/cgi/ir_submit.cgi?context=som_pub We attached the file that will be upload in the UTRGV repository

## Acknowledgments

Preliminary results of this study were presented and recognized as the best oral clinical presentation at the UTRGV School of Medicine’s 5 th Annual Research Symposium, 2021, Mission, TX. The study was supported by Convocatoria 2021-01: Impulso a la Investigación Científica y de Tecnología Aplicada, COTACyT grant number: COTACYT-2021-01-23. The authors acknowledge the generous support provided by the Universidad Mexico Americana del Norte, the Universidad Autónoma de Tamaulipas for supporting ESFUERSO sharing spaces, personnel, laboratory facilities, and the enthusiastic participation of alumni, staff, and faculty.

